# The consequences of adjustment, correction and selection in genome-wide association studies used for two-sample Mendelian randomization

**DOI:** 10.1101/2020.07.13.20152413

**Authors:** Venexia M Walker, Sean Harrison, Alice R Carter, Dipender Gill, Ioanna Tzoulaki, Neil M Davies

**Author notes:** Correspondence to: Venexia Walker; Bristol Medical School, University of Bristol, Oakfield House, Oakfield Grove, Bristol, BS8 2BN.

## Abstract

**Introduction:** Genome-wide association studies (GWASs) often adjust for covariates, correct for medication use, or select on medication users. If these summary statistics are used in two-sample Mendelian randomization analyses, estimates may be biased. We used simulations to investigate how GWAS adjustment, correction and selection affects these estimates and performed an analysis in UK Biobank to provide an empirical example.

**Methods:** We simulated six GWASs: no adjustment for a covariate, correction for medication use, or selection on medication users; adjustment only; selection only; correction only; both adjustment and selection; and both adjustment and correction. We then ran two-sample Mendelian randomization analyses using these GWASs to evaluate bias. We also performed equivalent GWASs using empirical data from 306,560 participants in UK Biobank with systolic blood pressure as the exposure and body mass index as the covariate and ran two-sample Mendelian randomization with coronary heart disease as the outcome.

**Results:** The simulation showed that estimates from GWASs with selection can produce biased two-sample Mendelian randomization estimates. Yet, we observed relatively little difference between empirical estimates of the effect of systolic blood pressure on coronary artery disease across the six scenarios.

**Conclusions:** Given the potential for bias from using GWASs with selection on Mendelian randomization estimates demonstrated in our simulation, careful consideration before using this approach is warranted. However, based on our empirical results, using adjusted, corrected or selected GWASs is unlikely to make a large difference to two-sample Mendelian randomization estimates in practice.

## INTRODUCTION

Concurrent with the advent of large-scale biobank studies collecting a vast array of genetic and phenotypic data, there has been a rapid increase in genome-wide association studies (GWASs) aiming to identify the genetic causes of common diseases or risk factors. Two-sample Mendelian randomization is a popular causal inference method that uses summary statistics from two GWASs to assess the effect of an exposure, obtained from one GWAS, on an outcome, obtained from the second GWAS. [1–3] It is a form of instrumental variable analysis and therefore requires four assumptions in order to obtain a point estimate for the causal effect, as summarised in Figure 1. Key advantages of two-sample Mendelian randomization include that it removes the need for individual level data and sample sizes can be maximized, particularly for rare outcomes, by using two separate studies instead of a single study that captures both the exposure and outcome.

**Figure 1.**
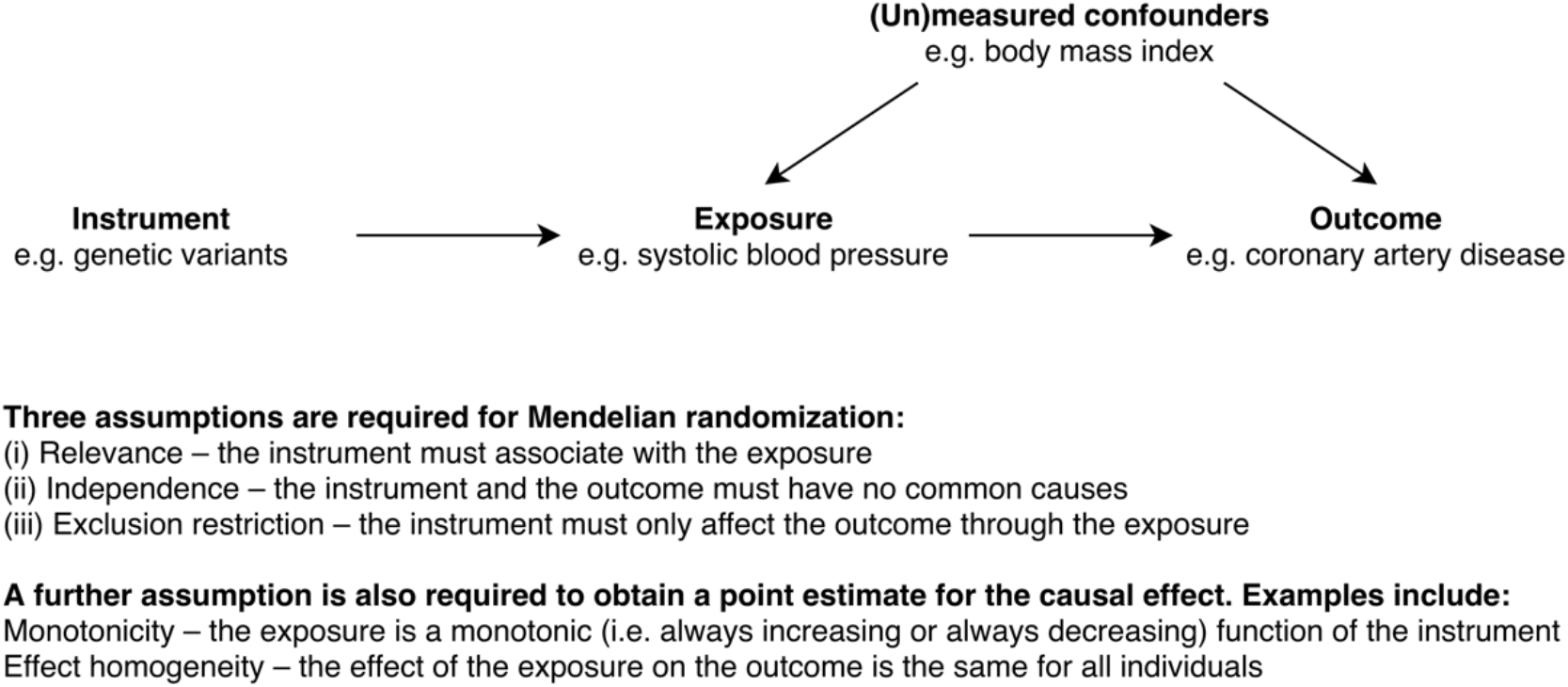
Schematic of Mendelian Randomization and the assumptions that must be satisfied for the results to be valid

Many GWASs used for two-sample Mendelian randomization analyses were conducted for other purposes, such as identifying individual single nucleotide polymorphisms (SNPs) associated with a phenotype. Consequently, GWASs are often adjusted for covariates, corrected for medication use, or selected on medication users to maximise power and avoid potential biases from factors that are not the phenotype of interest. However, while these practices are often beneficial for GWASs, they may affect the SNP-phenotype associations and consequently may cause bias in two-sample Mendelian randomization analyses. [4] The implications of adjustment for covariates in GWASs has been previously discussed, yet the consequences of correcting and selecting for treatment have not been fully described. [3,5,6] The aim of this study was therefore to assess the consequences of three common GWAS alterations and, where appropriate, their combination on two-sample Mendelian randomization estimates:

- Adjusting for a covariate – for example, including body mass index as a covariate in a GWAS of systolic blood pressure
- Correcting for medication use – for example, adding 10 mmHg to systolic blood pressure measures of individuals exposed to antihypertensive medicine prior to their blood pressure measurement [7]
- Selecting on medication users – for example, removing individuals receiving antihypertensive medicine prior to their blood pressure measurement

## METHODS

To demonstrate the possible consequences of using adjusted, corrected or selected GWASs in two-sample Mendelian randomization, we simulated genetic associations for six scenarios. These were: 1) no adjustment, correction or selection; 2) adjustment for a covariate; 3) selection on medication users; 4) correction for medication use; 5) adjustment for a covariate and selection on medication users; and 6) adjustment for a covariate and correction for medication use. These scenarios are illustrated using directed acyclic graphs in Figure 2. We then performed two-sample Mendelian randomization using the genetic associations from each of the scenarios and compared the results with the simulated effect of the exposure on the outcome. To demonstrate these consequences in a more realistic setting, we also conducted a comparable analysis using empirical data from UK Biobank with systolic blood pressure as the exposure, body mass index as the covariate and coronary heart disease as the outcome. [8]

**Figure 2.**
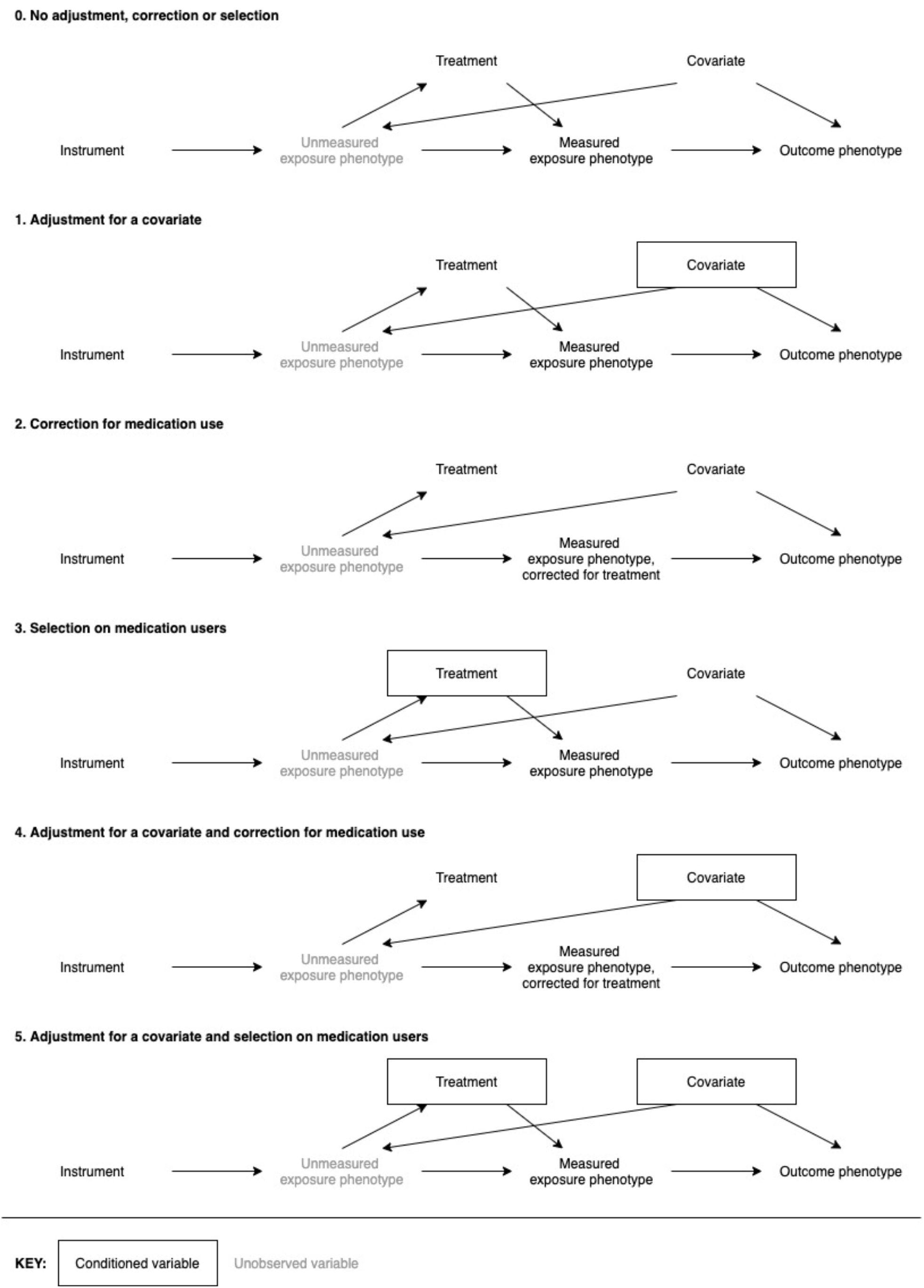
Directed acyclic graphs illustrating the six models considered in this study

**Figure 3.**
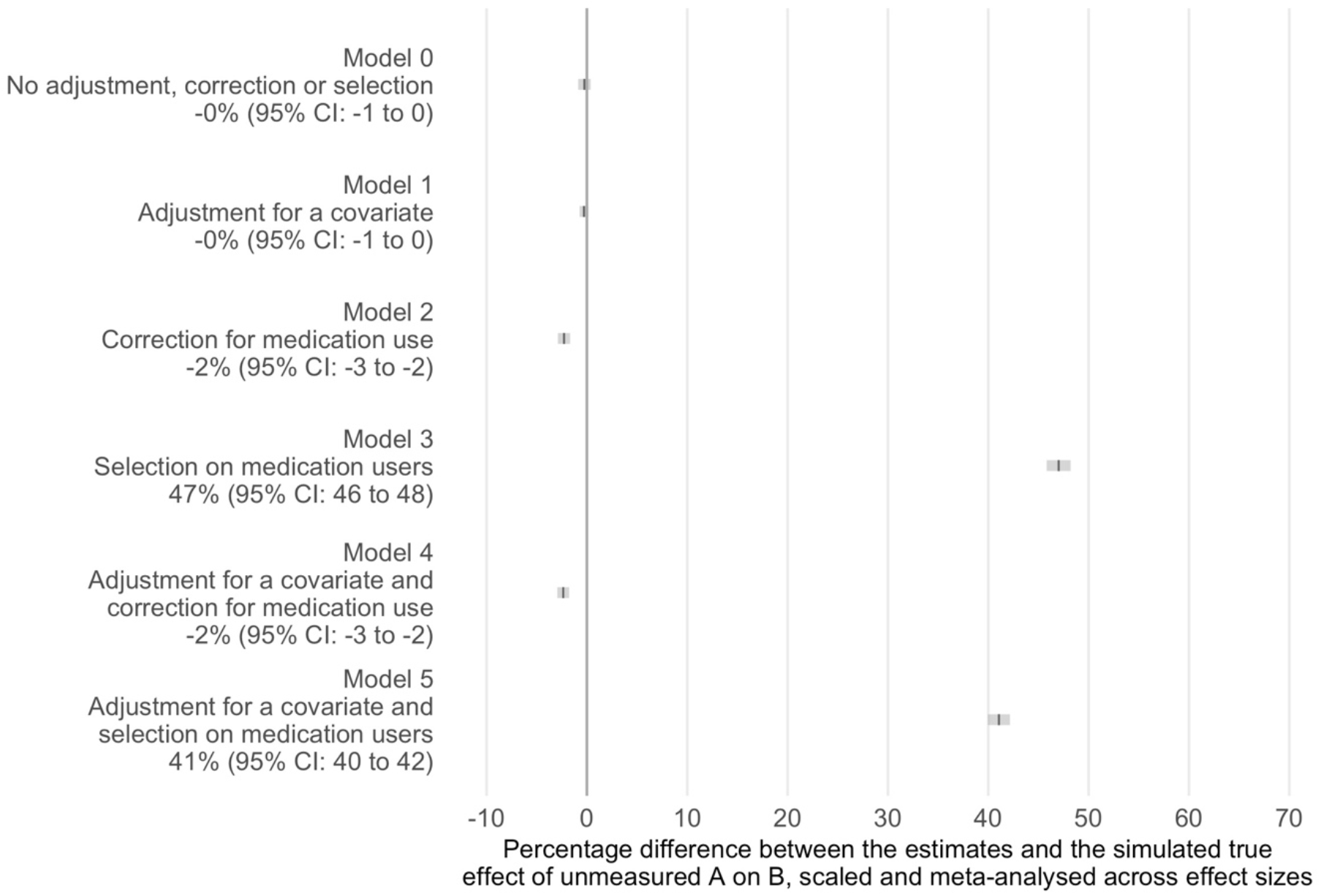
Percentage differences between two-sample Mendelian randomization estimates and the simulated true effect of unmeasured exposure phenotype on the outcome phenotype for different instrument-exposure association models

**Figure 4.**
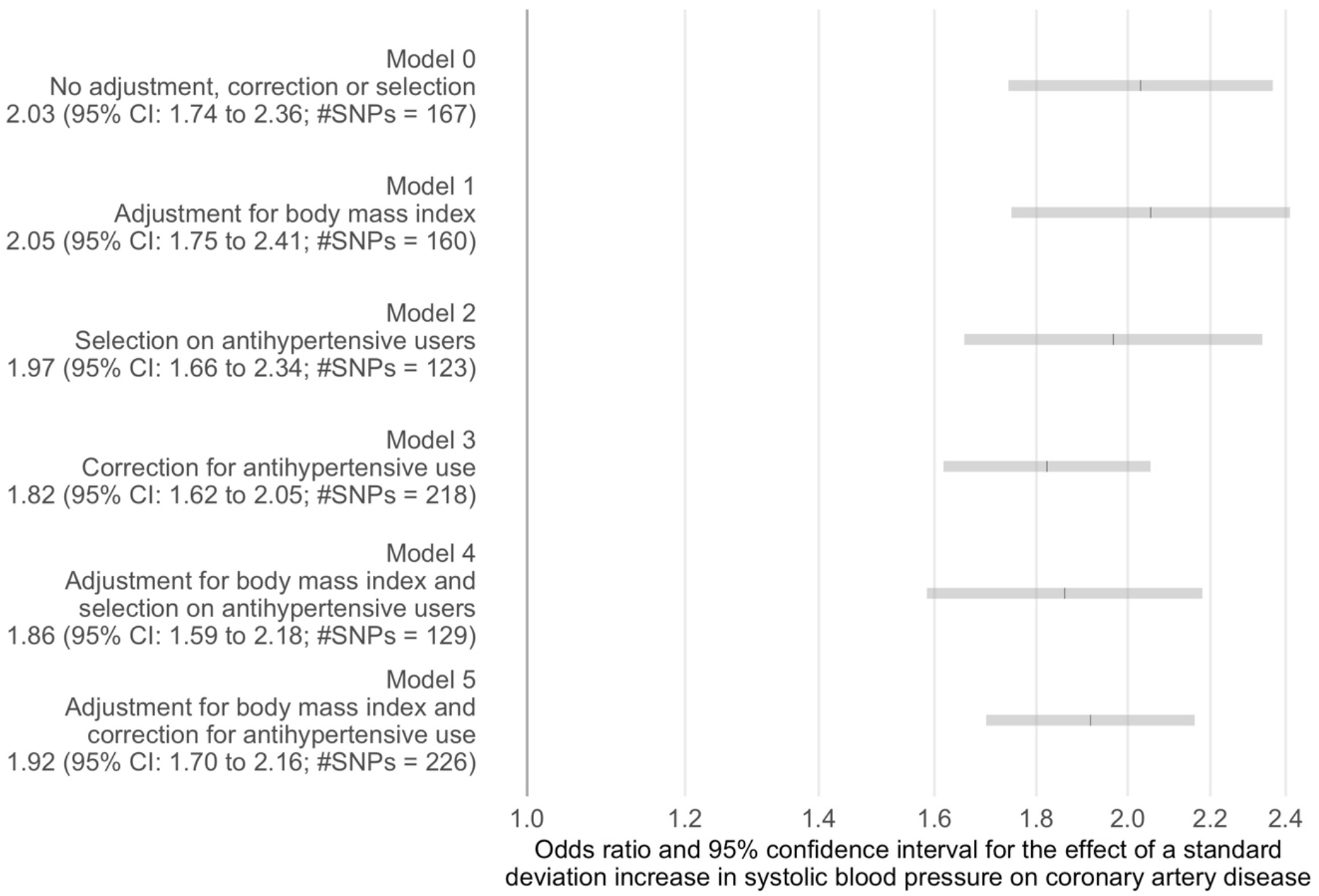
Two-sample Mendelian randomization estimates of the effect of systolic blood pressure on coronary artery disease for different instrument-systolic blood pressure association models

### Simulation

#### Data generation

We conducted a simulation based on a sample of 1,000,000 individuals with 200 SNPs that instrument our exposure. The simulation included specification of six parameters:

*α*_*CA*_, *α*_*AT*_, *α*_*AA\**_, *α*_*TA\**_, *α*_*CB*_ and *α*_*A*B*_. We simulated the genetic variants as a binomial function to get a SNP dose multiplied by a random effect of the SNP on the exposure phenotype. The formula for this, and the other main components of the simulation, are detailed below.

- Genetic variants, *Z*_*i*_ = Binomial(2, Uniform(0,1)) × Normal(0,0.1) for 1 ≤ *i* ≤ 200
- A continuous covariate, *C* = Normal(0,1)
- Unmeasured exposure phenotype, *A* = ∑_*i*_ *Z*_*i*_ + *α*_*CA*_*C* + *ε*_*A*_ where *α*_*CA*_ = 1, and *ε*_*A*_ = Normal(0, 2.5) is a random error term
- A treatment indicator, denoted by *T* = 1 if *p*_*t*_> Uniform(0,1) and 0 otherwise where 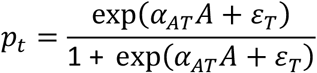 and *ε*_*T*_ = Normal(0, 1) is a random error term
- Measured exposure phenotype, *A\** = *α*_*AA\**_*A* + *α*_*TA\**_*T* + *ε*_*A\**_ where *α*_*AA\**_ = 1, *α*_*TA\**_ = −0.2, and *ε*_*A\**_ = Normal(0, 1) is a random error term
- Outcome phenotype: *B* = *α*_*A*B*_*A\** + *α*_*CB*_*C* + *ε*_*B*_ where *α*_*A*B*_ =various. *α*_*CB*_ = 1, and *ε*_*B*_ = Normal(0, 1) is a random error term

Genetic associations for the exposure phenotype

To obtain genetic associations for the exposure phenotype, we performed the following for a random sample of half the simulated individuals:

(0) Regressed the measured exposure phenotype on all genetic variants
(1) Regressed the measured exposure phenotype on all genetic variants, adjusting for the covariate
(2) Regressed the measured exposure phenotype on all genetic variants, excluding those indicated for treatment (*T* = 1)
(3) Regressed the corrected, measured exposure phenotype on all genetic variants – the correction was made by subtracting the product of the treatment indicator and the coefficient for the treatment indicator T from the regression *A\**~ *T* + *A* from the measured exposure phenotype value
(4) Regressed measured exposure phenotype on all genetic variants, adjusting for the covariate and excluding those indicated for treatment (*T* = 1)
(5) Regressed the corrected, measured exposure phenotype on all genetic variants, adjusting for the covariate – the correction was made by subtracting the product of the treatment indicator and the coefficient for the treatment indicator T from the regression *A\**~ *T* + *A* from the measured exposure phenotype value

#### Genetic associations for the outcome phenotype

We obtained genetic associations for the outcome phenotype by regressing the outcome phenotype on all genetic variants for the individuals that were not used to calculate the genetic associations for the exposure phenotype. This prevented bias from sample overlap. [9]

#### Mendelian randomization analysis and comparison with true effect

The true effect of the unmeasured exposure phenotype on the outcome phenotype per unit increase in the exposure in the simulation is equal to *α*_*A* B*_ *α*_*AA\**_. We simulated data for values of *α*_*A*B*_ between −1 and 1 in increments of 0.1, excluding zero. We performed Mendelian randomization for each dataset using the ‘mrrobust’ package in Stata. [10] We then calculated the difference between the Mendelian randomization estimate and the value of *α*_*A*B*_ *α*_*AA\**_. To ensure the effects were comparable, despite the different true effects, we scaled the differences by dividing by the value of *α*_*A* B*_ *α*_*AA\**_. Finally, we meta-analysed these scaled differences across all 20 simulated datasets using the *‘*metan*’* package in Stata with a fixed effect model and Mantel-Haenszel method. [11]

### Empirical analysis

#### Data source

We conducted the systolic blood pressure GWAS in UK Biobank. [8,12] UK Biobank recruited 503,317 UK adults between 2006 and 2010, collecting a wealth of phenotypic and genetic data. Analyses for this study were carried out on unrelated individuals of White British ancestry with a systolic blood pressure measure (N = 306,560, including 67,727 antihypertensive users). Individuals reported their ancestry, which was confirmed by assessing the consistency of individuals*’* genetic principal components with those of a European reference panel computed from the 1000 genomes project that was derived by UK Biobank. [13]

#### Genetic associations for systolic blood pressure

We used the mean of two systolic blood pressure readings, which were automatically recorded two minutes apart for all participants at the baseline UK Biobank assessment centre, to determine systolic blood pressure. Body mass index was calculated at the baseline assessment centre from baseline measures of height and weight. A list of antihypertensive medication was defined based on twelve headings from the British National Formulary (*adrenergic neuron blocking drugs; alpha-adrenoceptor blockers, angiotensin-converting enzyme inhibitors, angiotensin-II receptor blockers, beta-adrenoceptor blockers, calcium channel blockers, centrally acting antihypertensive drugs, loop diuretics, potassium-sparing diuretics and aldosterone antagonists, renin inhibitors, thiazides and related diuretics*, and *vasodilator antihypertensives*). [14] Antihypertensive medication use was then determined based on this list from medication records made by clinic nurses at baseline.

To obtain the SNP-systolic blood pressure associations for each scenario, we did the following:

(0) Performed a GWAS of systolic blood pressure in all individuals
(1) Performed a GWAS of systolic blood pressure in all individuals with body mass index included as a covariate
(2) Added 10 mmHg to systolic blood pressure for individuals who reported antihypertensive medication use at baseline and performed a GWAS of systolic blood pressure in all individuals
(3) Removed individuals who reported antihypertensive medication use at baseline and performed a GWAS of systolic blood pressure in the remaining individuals
(4) Added 10 mmHg to systolic blood pressure for individuals who reported antihypertensive medication use at baseline and performed a GWAS of systolic blood pressure in all individuals with body mass index included as a covariate
(5) Removed individuals who reported antihypertensive medication use at baseline and performed a GWAS of systolic blood pressure in the remaining individuals with body mass index included as a covariate

The value of 10 mmHg was chosen as the correction for antihypertensive medication use based on the fact that antihypertensives are estimated to reduce systolic blood pressure by 9 mmHg on average. [7] All GWASs were conducted using the MRC Integrative Epidemiology Unit Pipeline with a BOLT-LMM model to account for population stratification. [15] All six GWASs were adjusted for age, sex and 40 principal components.

#### Genetic associations for coronary artery disease

We obtained genetic associations for coronary artery disease from a multi-ethnic meta-analysis study by Nikpay et al of 60,801 cases and 123,504 controls to avoid sample overlap with UK Biobank. [16] This study used an inclusive definition for disease cases including clinically documented cardiac angina, coronary artery stenosis greater than 50%, and coronary revascularization.

#### Mendelian randomization analysis

Genome-wide significant hits from each GWAS were clumped using the *‘*clump_data*’* command in the *‘*TwoSampleMR*’* R package with a R squared threshold of 0.001 and a clumping window of 10,000kb. [17] These SNPs were then used as the instrument for the two-sample Mendelian randomization analysis, also performed using the *‘*TwoSampleMR*’* R package. All estimates were scaled to the effect per standard deviation increase in systolic blood pressure for presentation.

### Code availability

All analyses were conducted in Stata version 15.1 and R version 3.4 or higher. The code is available from GitHub: https://github.com/venexia/MR-GWAS-consequences.

## RESULTS

### Simulation

We found selection on medication users led to a point estimate of the beta that was 47% (95% CI: 46 to 48) greater than the simulated true effect of unmeasured exposure phenotype on the outcome phenotype across effect sizes. The combination of adjustment for a covariate and selection on medication users also led to an overestimate of the beta of 41% (95% CI: 40 to 42). There was little difference between the estimates and the simulated true effect of unmeasured exposure phenotype on the outcome phenotype obtained in the other scenarios.

### Empirical analysis

The estimates of the odds ratio for a standard deviation increase in systolic blood pressure on coronary artery disease varied between 1.82 (95% CI: 1.62 to 2.05; #SNPs = 218) for the model corrected for antihypertensive use only to 2.05 (95% CI: 1.75 to 2.41; #SNPs = 160) for the model adjusted for body mass index only. The confidence intervals for all estimates were overlapping suggesting little difference between estimates based on the different exposure GWAS used for these analyses.

## DISCUSSION

The aim of this study was to assess the consequences of using adjusted, corrected or selected GWASs on two-sample Mendelian randomization estimates. We demonstrated through simulations that selection of medication users can lead to substantial bias in Mendelian randomization estimates. On the other hand, bias from adjustment for a covariate and correction for medication use was minimal in the simulated scenarios. We have also presented results from an empirical analysis, where we used Mendelian randomization to estimate the effect of systolic blood pressure on coronary artery disease with body mass index as the covariate. We found little difference in the estimates for the effect of a standard deviation increase in systolic blood pressure on coronary artery disease across the six scenarios, even those selecting on medication users. Furthermore, in this example, the observed differences would not have affected the inference drawn from the analysis. This suggests that the bias observed in the simulations may be less pronounced in practice.

There are several potential causes of bias in the six scenarios we consider (Figure 2) that may explain our simulated results. In the case of scenario 0 (no adjustment, correction or selection) and scenario 1 (adjustment for a covariate only), ignoring the effect of treatment can lead to model misspecification. This is because the relationship between the instrument and the exposure will be a mixture of two distributions – one for those who are treated and one for those who are untreated. If the effect of the treatment on the exposure is large, the simple linear model used to capture the relationship between the instrument and the exposure will be inadequate to describe the mixture of these two distributions. In the remaining scenarios, where treatment is accounted for through selection on medication users or by adjusting for treatment effect, treatment becomes a collider and can introduce collider bias. This is because accounting for treatment in these ways is akin to conditioning on treatment and can therefore introduce spurious associations between any confounders and the genetic variants. This, in turn, leads to incorrect estimation of the effect of the genetic variants on the exposure. When these effects are then used in two-sample Mendelian randomization analyses, they may lead to bias in the estimation of the exposure-outcome effect.

The potential for bias in two-sample Mendelian randomization estimates using GWASs with adjustment, correction or selection has previously been highlighted as a limitation of this method in the literature. [18] For instance, Hartwig et al investigated the impact of using adjusted GWASs on two-sample Mendelian randomization estimates using a similar approach. [5] There has also been detailed discussion of the impact of selection and the potential for collider bias. For example, the idea of selection on medication use presented here relates to the selection criterion previously discussed by Gkatzionis and Burgess. [6] However, to our knowledge, there are no previous studies that have considered how the use of GWASs with a combination of adjustment for a covariate and measures to account for treatment, such as correction or selection, may impact two-sample Mendelian randomization estimates. This is despite several examples, including blood pressure, where such combinations are commonplace. Future work in this area should look to consider the potential for bias from adjustment, correction and selection in other implementations of Mendelian randomization, such as multivariable Mendelian randomization. [19]

While knowing the data generation mechanism for a simulation is an advantage, generated data is a simplification of real data generation mechanisms. Reality is likely much more complicated than the models we have studied. Our findings may therefore be considered indicative of potential bias in the scenarios we have outlined, rather than a quantification of the potential bias in all possible scenarios. For example, correction for medication use may have had little effect here because treatment occurs between the instrument and measured exposure phenotype, without having a direct effect on the outcome phenotype, and so should be captured within the GWAS of measured exposure phenotype. A further consideration is that our empirical analyses may also be subject to the common limitations of Mendelian randomization, such as horizontal pleiotropy. This may serve to amplify or disguise differences between estimates due to adjustment, correction and/or selection in the exposure GWAS.

Using simulations, we have shown that bias is possible in two-sample Mendelian randomization when using GWASs with selection on medication users. Careful consideration is therefore warranted before using this approach to account for treatment. Despite this, little difference was observed between the two-sample Mendelian randomization estimates in our empirical example and the inference drawn from these analyses remained the same for all six scenarios studied. This suggests that, in practice, using adjusted, corrected or selected GWASs is unlikely to make a large difference to estimates obtained from two-sample Mendelian randomization.

## Data Availability

Empirical data is available on application from UK Biobank. Simulated data is available from GitHub: https://github.com/venexia/MR-GWAS-consequences.

## FUNDING

VMW, ARC, SH and NMD are members of the Medical Research Council Integrative Epidemiology Unit, which is supported by the Medical Research Council and the University of Bristol [MC_UU_00011/1 and MC_UU_00011/4]. DG is supported by the Wellcome Trust 4i Programme (203928/Z/16/Z) and British Heart Foundation Centre of Research Excellence (RE/18/4/34215) at Imperial College London. NMD is also supported by a Norwegian Research Council [grant number 295989].

## CONFLICTS OF INTEREST

DG is employed part-time by Novo Nordisk.

## DATA AVAILABILITY

Empirical data is available on application from UK Biobank. The presented research has been conducted under Application Number 16729. Simulated data is available from GitHub.

## AUTHOR CONTRIBUTIONS

VMW and SH performed the simulations. ARC and SH performed the GWAS analysis using the empirical data. DG and VMW performed the Mendelian randomization analysis using the empirical data. VMW drafted the manuscript. All authors interpreted the results and revised the manuscript.

## ACKNOWLEDGEMENTS

Quality Control filtering of the UK Biobank data was conducted by R.Mitchell, G.Hemani, T.Dudding, L.Paternoster as described in the published protocol (doi:10.5523/bris.3074krb6t2frj29yh2b03×3wxj). The MRC IEU UK Biobank GWAS pipeline was developed by B.Elsworth, R.Mitchell, C.Raistrick, L.Paternoster, G.Hemani, T.Gaunt (doi: 10.5523/bris.pnoat8cxo0u52p6ynfaekeigi).

## Notes

### Clinical Trial

This research has been conducted using the UK Biobank Resource under Application Number 16729.

### Author Declarations

This research has been conducted using the UK Biobank Resource under Application Number 16729.

